# Does time awareness coaching support hybrid workers’ wellbeing?: Protocol for a pilot randomized controlled trial

**DOI:** 10.1101/2024.06.28.24309658

**Authors:** Anna Navin Young, Zelda Di Blasi, Sarah Foley, Eithne Hunt

**Affiliations:** School of Applied Psychology, University College Cork, Ireland; Department of Occupational Science and Occupational Therapy, University College Cork, Ireland

## Abstract

**Background:** High rates of poor employee mental health and wellbeing have spurred growing demands for initiatives that support wellbeing in the workplace. The promotion of positive mental health and wellbeing is an essential component of workplace wellbeing initiatives, focusing on enhancing positive aspects of work, workers’ capacities, and positive behaviors. As one of the fastest growing practices in personal and professional development, coaching is found to improve workers’ wellbeing and performance through reflection, awareness, and meaningful goal pursuit. As time-related challenges exacerbate workplace stressors and threaten wellbeing, specific time-focused coaching approaches are essential to the promotion of positive mental health and wellbeing in the workplace. Effectively addressing work-time challenges is especially critical for hybrid workers, who divide their work-time across multiple onsite and remote locations.

**Methods:** The current paper is a protocol for a pilot three-armed randomized controlled trial that aims to evaluate the effectiveness of a time awareness coaching (TAC) intervention to support hybrid workers’ wellbeing. Sixty hybrid working participants will be randomized to either the TAC intervention, reflective time tracking (active control), or a passive control group. Professional coaches will be recruited and trained to facilitate the TAC intervention. Pre-post intervention measures of chronic time pressure, perceived control of time, perceived stress, wellbeing, and self-efficacy will be evaluated and compared across intervention arms. Qualitative feedback from participants and coaches will be collected to assess the intervention’s acceptability and implementation.

**Discussion:** The results of the study will offer insights into intervention effectiveness as well as the feasibility of critical intervention elements such as recruitment, retention, and implementation. The findings will inform recommendations for the use of TAC in future research studies and workplace wellbeing initiatives.

## Introduction

Beyond its impacts on absenteeism, job retention, impaired quality of life, and decreased productivity and performance, poor mental health is a leading cost in worker compensation systems and is estimated to cost the global economy $1 trillion annually [1–4]. The World Health Organization [5] defines mental health as, “a state of mental well-being that enables people to cope with the stresses of life, realize their abilities, learn well and work well, and contribute to their community”. In recent years, workplace mental health and wellbeing has become a billion-dollar industry populated with an array of unregulated offerings [1,7]. As a result, various governments and global organizations have sought to develop policy and practice guidelines to help regulate and facilitate consistent and effective initiatives across workplaces [3,7,8]. Thus, it is essential that mental health and wellbeing services are validated and evidence-based to ensure they can be effectively implemented.

A recent umbrella review of workplace mental health interventions identified three main types of interventions: protection from harm, promotion of mental health and wellbeing, and addressing needs of those at increased risk [1]. The promotion of mental health and wellbeing has gained increasing attention in past decades, with promising results for individuals and organizations [9–11]. Interventions that aim to promote mental health and wellbeing typically focus on developing positive aspects of work, positive worker capacities, and positive behaviors [6, 12–15]. By focusing on factors such as employee strengths, resilience, participatory job crafting, and intentional work habits, these promotion-focused interventions aim to enhance positive outcomes such as wellbeing, engagement, self-efficacy, and optimal functioning [1, 9, 12]. Furthermore, the positive outcomes that arise from promotion-based interventions can in turn buffer against negative mental health consequences [9, 16].

As one of the fastest growing approaches to learning and development within organizations [17], coaching has great potential as an evidence-based initiative to promote mental health and wellbeing in the workplace. Across industries, coaching is already a widely implemented approach to supporting employee wellbeing and performance [18]. Previous meta-analyses evaluating the effectiveness of coaching in the workplace found positive impacts on a wide array of wellbeing and performance outcomes [19–21].

Rather than taking a prescriptive approach to defining program success, coaching is fundamentally a person-centered approach [22, 23]. As a person-centered approach, coaching places emphasis on the coachee’s active role in shaping the coaching experience, seeing personal values, experiences, and goals as core drivers of the coaching work [23, 24]. This person-centered approach prompts positive outcomes such as enhanced self-regulation and self-efficacy through self-reflection, self-awareness, and positive behavior change [19, 20, 23].

### Time Awareness Coaching

Time-focused initiatives are an essential component of supporting wellbeing in the workplace. How workers objectively use and subjectively experience their time have important implications for their wellbeing [25–27]. Work intensification is a well-studied example of this, characterized by a faster pace of work, tighter deadlines, a reduction in idle and undisturbed time, and instances of engaging with multiple work tasks simultaneously [28]. These work intensification characteristics are enacted both through time use (e.g., multitasking) and perceptions of time (e.g., faster pace) and may involve feeling chronically pressed for time and out of control of time [26, 29, 30]. These patterns in time use and subjective experiences of time are becoming widely recognized as an occupational hazard and an important psychosocial risk in the workplace due to their impacts on worker wellbeing, mental health, motivation, performance, and functioning [29, 31].

Time researchers suggest that building awareness of current time habits and their impacts is a critical step to becoming more intentional and in control of our time [25, 27]. Using reflection and self-awareness, coaching can support coachees to recognize their current work-time habits and how they may be helping or hindering their wellbeing. Additionally, coaching is designed to support personally meaningful behavior change, thus enabling workers to apply their insights as appropriate to enhance their wellbeing at work. A coaching approach specifically designed to facilitate a coachee’s awareness of their time habits in connection to their personal and professional goals, values, experiences, and wellbeing may thus serve as a valuable initiative in the promotion of mental health and wellbeing in the workplace. Previous studies have suggested the usefulness of the time awareness coaching approach, [32, 33], however further evidence is needed to evaluate its effectiveness in a workplace context.

#### Hybrid Workers

The topic of time-use and subjective experiences is particularly relevant to address in the shifting landscape of office-based work. Hybrid work, for example, has become increasingly common and desirable since the COVID-19 pandemic expedited organizations’ adoption of nontraditional work models [34]. Hybrid work refers to a role where work time is divided between in-person (e.g., onsite) and remote (e.g., home) locations [35].

The hybrid work model reflects an increased flexibility of spatial and temporal work characteristics, and available literature indicates that efficient time and attention management are critical predictors of job performance and affective outcomes in hybrid workers [34]. Though the available literature is still scarce, it appears that work-time practices that enable workers to autonomously manage their time, set and maintain work-time boundaries, and effectively manage distractions and interruptions will play critical roles in enabling workers to work well and sustain their wellbeing while hybrid working [34–36]. Thus, the current study aims to evaluate whether a time awareness coaching intervention can effectively support hybrid workers’ work-time practices and wellbeing.

## Materials and methods

### Study aims and design

The study is a pilot three-arm pilot randomized controlled trial consisting of an experimental group (time awareness coaching; TAC), an active control group (reflective time journaling), and a passive control group. The trial is designed to evaluate the effectiveness of the TAC intervention in comparison with the active and passive control groups among a population of hybrid workers. Participant outcomes will be assessed pre- and post-intervention to evaluate chronic time pressure, perceived control of time, self-efficacy, perceived stress, and wellbeing. The reporting of this protocol has been informed by the SPIRIT checklist (Standard Protocol Items: Recommendations for Interventional Trials; see S1 Checklist) [37].

#### Inclusion criteria

Individuals will be eligible to participate if they are (1) 18 years or older, (2) currently work in a hybrid work environment, and (3) are English speaking. In this study, hybrid work is classified as a position someone fulfills whilst dividing their work time between in-person (office) and remote (e.g. at home) environments [35]. Using convenience and snowball sampling, authors will recruit participants through social media platforms (e.g. LinkedIn) and through recruitment calls via their professional networks.

#### Exclusion criteria

Anyone working in a fully remote or fully onsite role will be ineligible to participate in this study. People experiencing severe mental health conditions are excluded as the coaching intervention is not designed to address these conditions. To address this exclusion criteria in the study, we observe current involvement in work as an indicator of a lack of severe mental health condition [3, 5].

### Coach practitioner involvement

Coaching practitioners will be recruited and trained to facilitate the TAC intervention. Approximately 6-8 coaches will be recruited. Practitioners will be eligible to participate if they are members of a professional coaching body such as the Association for Coaching (AC), International Coach Federation (ICF), and the European Mentoring and Coaching Council (EMCC). Membership with a professional body indicates a standardized level of practitioner training, commitment to continued professional development (CPD), and adherence to ethical codes of conduct. Coaches will be recruited through social media platforms (e.g. LinkedIn) and through convenience and snowball sampling where authors share a recruitment call through their networks.

#### Practitioner training

Once coaches are recruited, they will be trained to facilitate the TAC intervention for this study. Training will involve a half-day virtual group training program facilitated by the first author, and the provision of client contact email templates and coaching session guidelines. The development of the training program has been informed by the procedures for TAC practitioner training [32], with specific tailoring for the trial context (e.g., overview of trial and context of intervention – hybrid work-time habits and wellbeing). Coaches will have ongoing contact with the first author to ensure they feel comfortable and confident adhering to the intervention procedures.

#### Practitioner data collection

After completing the training program, coaches will be instructed to complete a brief online survey to collect basic demographic information and background information regarding their coaching experience (see Figure 1). Demographic information such as age and gender will be collected, along with professional information such as number of years practicing as a coach and with which professional body they hold membership. This information will be summarized in the final trial write up. At the end of the trial when coaches have concluded their work with participants, they will again be instructed to complete a final online survey. This survey will ask them about their experiences of training in and facilitating the TAC intervention. Coaches will be asked whether they would use the TAC approach with their own clients outside of the study context, and whether they would recommend the approach to other coaching practitioners. This feedback will inform the study’s overall implications regarding future research and practice using the TAC approach.

**Fig 1.**
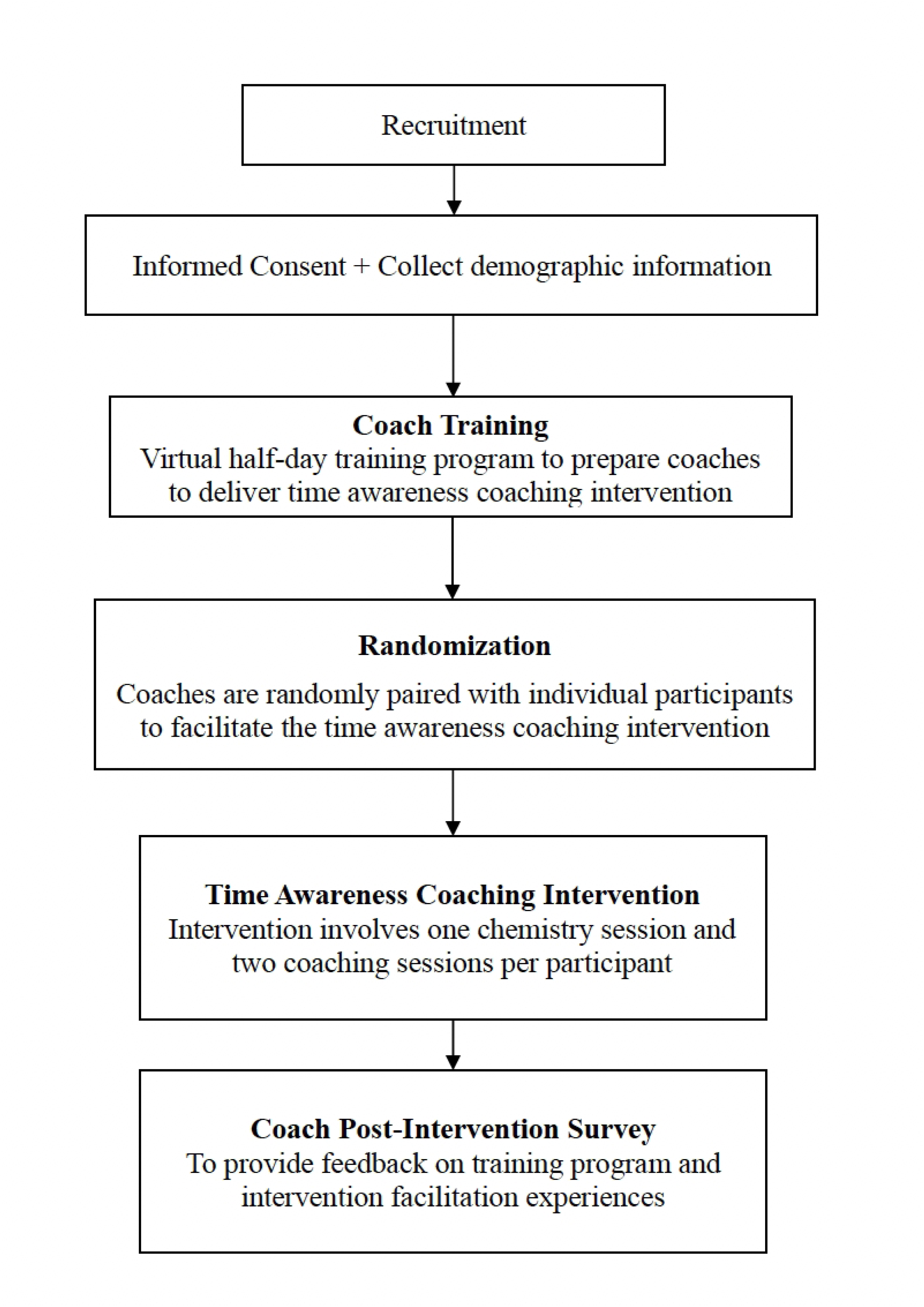
Flow chart of coach involvement.

### Procedures

Participation in this experiment involves three stages: (1) initial survey, (2) time awareness activity (three intervention arms), and (3) post-time awareness activity survey (see Figure 2). All participant involvement will take place online and virtually. Survey data collection will be facilitated using Qualtrics [38]. Participants will obtain access to the first survey after they have been presented with an information-consent form and indicated their consent. When participants complete the first survey, they are randomized to one of the three intervention arms and receive the relevant instructions. They are encouraged to contact the research team should they have any questions regarding the activity they are meant to complete (and are provided with contact information).

**Fig 2.**
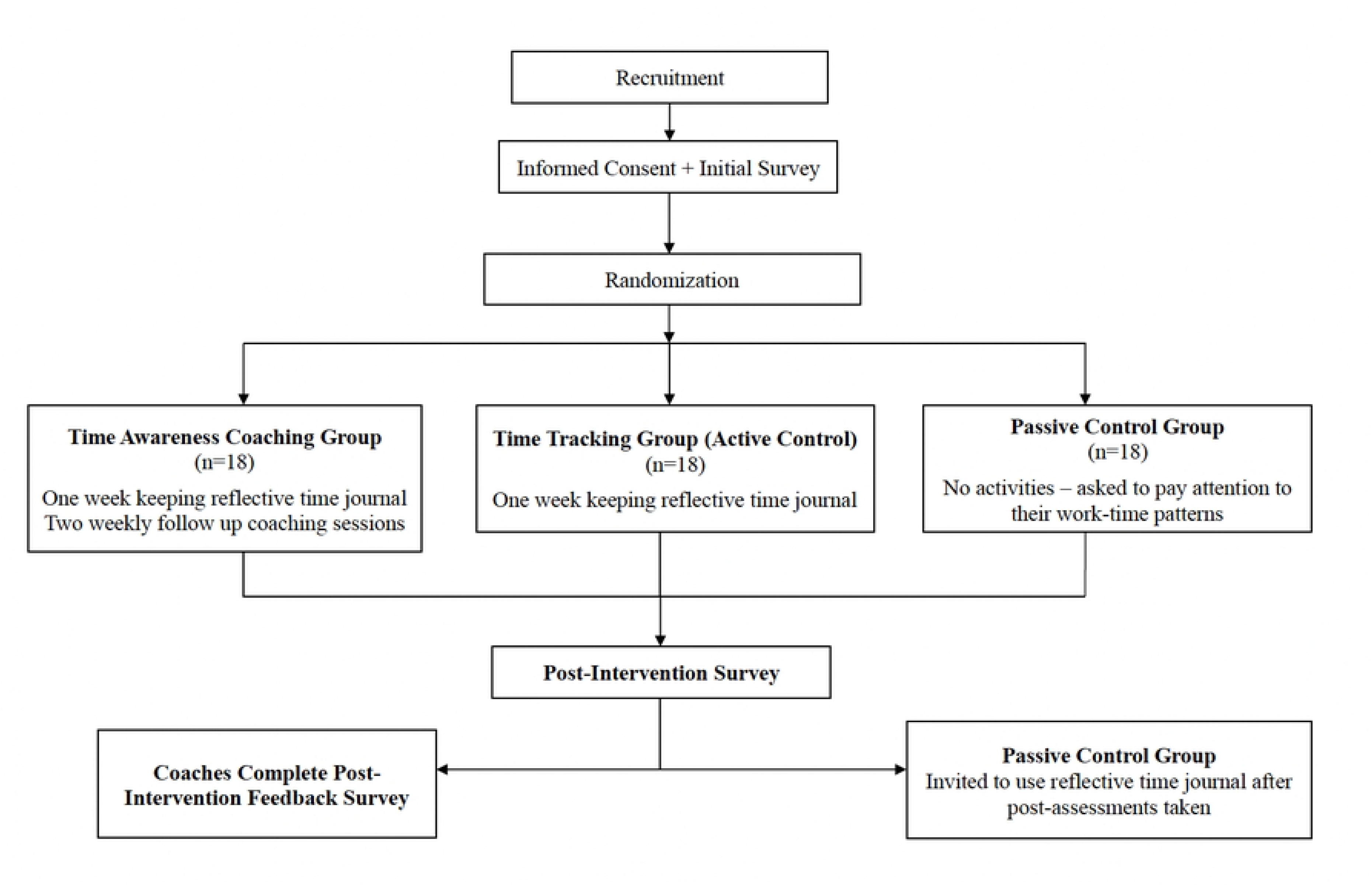
Flow chart of participant involvement.

At each stage of participant involvement, participants are provided with both text-based and video-based instructions. For example, when participants click into the first survey, the information-consent form is provided in text and is also read out by the first author in a short video. When participants receive email reminders to prompt them to continue with their intervention, the emails contain a brief written reminder as well as a link to a video reminder. With the aim of establishing consistency and supporting retention, the first author is the main subject of all video recordings, the videos remain visually consistent (same digital background), and participants are welcomed in each video with a reminder of the study’s purpose (to investigate hybrid workers’ work-time practices and wellbeing).

#### Randomization

When participants complete the first survey, they will be randomized to one of the three intervention arms using Qualtrics’ randomization features [38]. After completing the first survey and being randomized into their intervention arm, participants will then receive instructions for completing their relevant time awareness intervention. The first author will facilitate coach-participant pairings by polling coaches for availability, assigning numbers, and rolling a die to randomly select an available coach to pair with each new experimental participant.

### Interventions

#### Time Awareness Coaching (TAC; experimental group)

When randomized to the experimental group, participants receive instructions and materials to complete a reflective time tracking activity. Participants are introduced to the digital reflective time journal (see S1 Appendix) and instructed on how to track their work time using the six columns of the journal. Participants are also instructed to use the time journal to record any reflections, insights, and feelings that arise for them as they track their work time and consider any patterns they notice in how they use and experience their time.

The reflective time journal template is provided as a “view only” spreadsheet. On the first tab of the spreadsheet, participants are provided instructions on how to either copy or download the template in order to create an editable file. The template consists of seven additional tabs, each representing one day of the tracking period. Each tab contains the same six-columned time tracking table (see S1 Appendix). Through video instructions, participants are provided with an overview of the template and how to track their time in 30-minute increments over the seven-day tracking period. While tracking, participants are instructed to distinguish between main activities that occur in each 30-minute increment as opposed to other activities that also may occur (these could include interruptions and distractions, for example). Additional columns in the time journal encourage participants to document their location, and their subjective rating of quality of time (on a scale of 1-low quality to 10-high quality). Finally, participants are encouraged to use the notes sections throughout the template to record any insights, thoughts, and feelings that arise throughout the tracking process.

Participants are instructed to complete the reflective time journal for a period of seven days and receive two email reminders throughout the week to encourage them to continue with the activity. TAC participants are additionally advised that they will be paired with a coaching practitioner and will receive two virtual coaching sessions to support them to apply insights from the reflective time journal into meaningful changes in their lives. Participants are provided with the following overview to explain the coaching component of the intervention:

> *“In addition to completing the time journal, you have the opportunity to work with a coaching practitioner. The coaching space is an opportunity to reflect on your insights from the time journal and receive support from a trained professional to put these insights into meaningful action. Through two virtual coaching sessions, your coach will support you to reflect on your time habits, identify desired changes, and pursue these changes to improve your work time and wellbeing.”*

Participants are then informed that they will be paired with a coach, who will contact them to schedule their coaching sessions. Participants are also provided with a coaching agreement document that outlines the scope of the coaching relationship. Contracting is a critical component of coaching and serves to establish clear expectations for the professional service as well as build rapport between the coach and client [39].

When a coach is paired with a participant, they will first have a brief phone call, known as the chemistry session, which serves as an opportunity for the coach and participant to get to know each other, confirm the scope of the coaching relationship, and clarify any confusions or questions before proceeding with the coaching sessions. These initial conversations are seen as a critical component of developing a trusting and effective coaching relationship [39]. After the chemistry session, and after the participant has completed the reflective time journaling, the coach and participant schedule two coaching sessions, approximately one week apart. At the end of the second coaching session, the coach provides the participant with the link to the follow-up survey. After participants complete the follow-up survey, they receive a debriefing message regarding the nature of the trial and the intervention arms.

#### Reflective time journaling (active control group)

When randomized to the active control group, participants receive the same instructions and materials as the TAC group and are encouraged to complete the seven-day time tracking activity. Unlike the TAC group, those randomized to the active control group do not receive coaching as part of their intervention. Two automated email reminders are sent throughout the week of tracking. Participants receive another email at the end of the seven-day period with a link to the follow-up survey. A final email is sent ten days after the completion of the first survey, reminding participants to complete the follow-up survey. When participants have completed the follow-up survey, they receive a debriefing message outlining the nature of the trial and the three intervention arms.

#### “Think about your time” (passive control group)

Participants randomized to the passive control group will receive the following instructions:

> *“For the next week, please pay attention to your work-time habits. Pay attention to any patterns you start to notice in when, where, and how you work, and how you feel while working.”*

Participants receive two email reminders throughout the week, reminding them to pay attention to their work-time habits. At the end of the seven-day period, participants receive another email with a link to the follow-up survey. Ten days after completing the first survey, participants receive a final email with a reminder to complete the follow-up survey if they have not yet done so. After completing the follow-up survey, participants are then debriefed on the nature of the trial and the three intervention arms. They receive the instructions and materials for the reflective time journaling and are encouraged to complete this activity for their own benefit should they wish to do so.

### Measures and outcomes

The initial survey will collect demographic information from participants, including age, gender, living arrangements, family and relationship status, and highest level of education.

#### Hybrid work characteristics

Alongside the demographic characteristics, the initial survey will also collect information regarding participants’ hybrid work contexts and preferences. Information gathered will include average days per week working in-person and remotely, average weekly work hours when working in-person and remotely, physical work setup in in-person and remote environments, level of personal control over where they work, and level of satisfaction with their current hybrid work schedule.

To contribute to the growing hybrid work literature, open-ended questions will encourage participants to provide further detail regarding their hybrid work experiences and preferences. These questions ask about factors that influence participants’ preferences for working in-person and remotely, the greatest benefits and challenges of working in-person and remotely, and qualities of their ideal work environment. The aim of including these open-ended questions is to collect nuanced insight into participants’ experiences to supplement the interpretation of quantitative responses.

#### Outcome measures

Self-reported measures will be collected using the Chronic Time Pressure Inventory [40], the Perceived Control of Time Scale [41], the General Self-Efficacy Scale (GSE) [42], the Perceived Stress Scale (PSS) [43], and the WHO-5 Well-Being Index [44]. These measures will be repeated in both the initial and follow-up surveys.

#### Time pressure

Time pressure is measured using the 13-item Chronic Time Pressure Inventory [40]. Items are measured on a five-point Likert scale from 1 (strongly disagree) to 5 (strongly agree). The item scores can be added up for a total chronic time pressure score ranging from 13 to 65. Items can also be divided across two factors: ‘feeling harried’ and ‘cognitive awareness of time shortage’ and summed to reach sub-scores. An example item of ‘feeling harried’ is: ‘The days fly by without me ever getting everything done.’ An example item of ‘cognitive awareness of time shortage’ is: ‘There aren’t enough hours in the day.’ Cronbach’s alphas demonstrated good internal consistency for the total scale (α = 0.85), for the ‘feeling harried sub-scale (α = 0.795), and for the ‘cognitive awareness of time shortage’ sub-scale (α = 0.80) [40].

#### Perceived control of time

The Perceived Control of Time Scale is a 5-item scale that assesses how respondents perceive their ability to directly affect and manage their time [41]. The scale takes a work-specific perspective to control of time, with reference to work-time in four of the five scale items [41]. Items are measured on a 5-point Likert scale from 1 (do not agree at all) to 5 (completely agree). On a range of five to 25, a higher total score indicates a higher level of perceived control of time. Sample items include: ‘I feel in control of my time,’ and ‘I often have little control of what is happening at work.’ The Cronbach’s alpha was 0.73 [41].

#### Self-efficacy

Schwarzer and Jerusalem’s General Self-Efficacy Scale is a widely used scale that consists of 10 items ranked from 1 (not at all true) to 4 (exactly true) [42]. The scale aims to assess respondents’ belief in their ability to cope with adversity and perform new or difficult tasks [45]. Total scores range from 10 to 40, with a higher score indicating a higher level of self-efficacy. A sample item is: ‘When I am confronted with a problem, I can usually find several solutions.’ Cronbach alphas have ranged from 0.76 to 0.9 [42, 46].

#### Perceived stress

The Perceived Stress Scale (PSS) [43] is the most widely used instrument for measuring appraisals of stress. The instrument assesses how respondents perceive their lives as stressful, particularly in how unpredictable, uncontrollable, and overloaded they perceive their lives to be. The PSS is a 10-item scale with items scored from 0 (never) to 4 (very often). A sample item is: ‘In the last month, how often have you felt that you were unable to control the important things in your life?’ When added up, total scores range between 0 and 40, with scores between 0 and 13 indicating low perceived stress, scores between 14 and 26 indicating moderate stress, and scores between 27 and 40 indicating high stress. Cronbach’s alpha ranged from 0.84 to 0.86 in initial instrument testing [43].

#### Well-being

The WHO-5 Well-being Index is one of the most widely used measures to assess subjective well-being [47].The Index consists of five statements which are responded to from 0 (at no time) to 5 (all of the time) [44].The total raw score ranges from 0 to 25 and is then multiplied by four to represent a final well-being score. Higher scores represent higher well-being. National general population averages range from 54-70 [47]. A sample item on the index includes, “I have felt cheerful and in good spirits.” Cronbach’s alphas have ranged from 0.83 to 0.95 across various participant populations [48,49].

#### Post-intervention experiences

In the follow-up survey, participants in the experimental and active control groups will also receive open-ended questions asking about their experiences of their respective time awareness activity. In addition to asking them to share their overall experience, specific questions will ask participants to reflect on anything that surprised them, any changes they made to their time use in response to the intervention, and anything they learned through their involvement in the intervention.

### Sample size

The present pilot study aims to evaluate whether the TAC intervention enhances hybrid workers’ wellbeing, self-efficacy, and perceived control of time while diminishing their chronic time pressure and perceived stress. The study aims to compare the TAC intervention’s effectiveness to that of the active (reflective time tracking) and passive control groups.

An a priori power analysis was calculated using G*Power 3.1 [50, 51] to determine the required sample size. Assuming a power (1-β) of 90%, and a significance level (α) of 0.05, the sample size was determined as 18 participants per group (54 participants total). To account for potential attrition (rate set at 10%), the total sample size for the study was set at 60 participants.

### Data management plan

All data generated in this study will be securely stored on university-approved storage services using password-protected files. Only the research team (listed authors) will have access to the raw data and materials involved in this study. Each participant is instructed to generate a code at the beginning of the first survey. They will input this code again at the beginning of the follow-up survey to allow for pre/post analyses. At the beginning of the first survey, participants are asked whether they consent for their data to be used in future research. Should participants consent to future data use, only completely anonymized data will be made available in data repositories. These data management practices will adhere to FAIR guidelines [52].

### Data analysis

The primary objective of this intervention development pilot study is to investigate critical elements of the intervention (e.g., recruitment, retention, acceptability) and to provide transparent reporting of intervention design and implementation to promote replicability. Descriptive data will be presented on the feasibility of recruitment and retention. Acceptability will be examined through qualitative feedback from coaches and participants. Coaches will be asked to provide feedback on the acceptability of the training program, the TAC intervention within the trial, and how useful and practical they feel the TAC program would be for coaching clients beyond the trial context. In the post-survey, TAC participants will be asked about their experiences of the intervention using open-ended questions. The active control group will also be asked about their experiences of the time tracking intervention. As appropriate, reflexive thematic analysis will be used to analyze participants open-ended responses to better understand their experiences of the intervention [53, 54].

Analyses will adhere to ‘intention-to-treat’ procedures with the inclusion of data from all randomized participants [55, 56]. Descriptive and demographic data collected in the pre-survey will be reported to present an overview of participants and their hybrid work environments. We will conduct mixed between-within analyses of variance to evaluate the effect of the TAC intervention on participants’ wellbeing, perceived stress, self-efficacy, chronic time pressure, and perceived control of time, and to further compare the TAC intervention to the active and passive control groups.

### Ethics approval

The study has received approval from the University’s ethics committee [Log number 2023-101; full details will be provided after peer review]. Participation is voluntary and informed consent will be obtained. When recruited, coaches will provide written consent to a coaching contract, outlining their involvement and their maintenance of client confidentiality. When coaches and participants are paired in the experimental group, they will verbally agree to a coaching agreement outlining the scope of the coaching relationship.

### Status and timeline

In February 2024, coaches were recruited and trained to facilitate the time awareness coaching intervention. Participant recruitment began on February 14th, 2024. The pilot trial is currently ongoing.

## Discussion

Evidence-based interventions are needed to effectively address high rates of poor employee mental health and wellbeing [1–8]. Specifically, further evidence is needed in the area of sustainable wellbeing promotion [1,9,10]. As coaching emerges as a fast-growing and effective approach in the workplace, further evaluation of specific coaching interventions will provide guidance for future workplace wellbeing initiatives. The aim of the current pilot randomized controlled trial is to evaluate the effectiveness, feasibility, and acceptability of a time awareness coaching intervention to support hybrid workers’ work-time practices and wellbeing. Specifically, the pilot trial will evaluate the time awareness coaching in relation to participants’ chronic time pressure, perceived control of time, perceived stress, wellbeing, and self-efficacy. The study will further collect qualitative feedback from participants to assess their experiences of the intervention. Additionally, the pilot trial will collect feasibility and acceptability information from the coaches recruited and trained to facilitate the intervention within the trial. This input can further inform recommendations for the practical use of the intervention.

The pilot study is a necessary step to investigate the novel intervention and will provide robust foundations for future research and practice initiatives [57]. The three-arm comparator design is a strength of the current study as it provides a realistic evaluation of critical study elements such as recruitment, randomization, retention, and intervention implementation [57]. Furthermore, the presentation of the current protocol aims to promote transparency and replicability in accordance with best practices in intervention reporting [37, 58]. The pilot study’s findings regarding effectiveness, feasibility, and acceptability will play a critical role in informing the design and implementation of future large-scale studies while also providing preliminary suggestions for workplace wellbeing initiatives that wish to incorporate coaching approaches.

## Supporting Information

### S1 Checklist

SPIRIT Checklist - Standard Protocol Items: Recommendations for Interventional Trials.

### S1 Appendix

One day of reflective time journal template.

## Data Availability

No datasets were generated or analysed during the current study. All relevant data from this study will be made available upon study completion.

